# Shedding of infectious virus in hospitalized patients with coronavirus disease-2019 (COVID-19): duration and key determinants

**DOI:** 10.1101/2020.06.08.20125310

**Authors:** Jeroen J.A. van Kampen, David A.M.C. van de Vijver, Pieter L.A. Fraaij, Bart L. Haagmans, Mart M. Lamers, Nisreen Okba, Johannes P.C. van den Akker, Henrik Endeman, Diederik A.M.P.J. Gommers, Jan J. Cornelissen, Rogier A.S. Hoek, Menno M. van der Eerden, Dennis A. Hesselink, Herold J. Metselaar, Annelies Verbon, Jurriaan E.M. de Steenwinkel, Georgina I. Aron, Eric C.M. van Gorp, Sander van Boheemen, Jolanda C. Voermans, Charles A.B. Boucher, Richard Molenkamp, Marion P.G. Koopmans, Corine Geurtsvankessel, Annemiek A. van der Eijk

**Author notes:** **Corresponding author:** Jeroen J.A. van Kampen, MD, PhD, Department of Viroscience, Unit Clinical Virology, Erasmus MC, Rotterdam, The Netherlands. equal contribution (Marion P.G. Koopmans, Corine Geurtsvankessel, Annemiek A. van der Eijk).

## Abstract

**Background:** Long-term shedding of viral RNA in COVID-19 prevents timely discharge from the hospital or de-escalation of infection prevention and control practices. Key questions are the duration and determinants of infectious virus shedding. We assessed these questions using virus cultures of respiratory tract samples from hospitalized COVID-19 patients as a proxy for infectious virus shedding.

**Methods:** Clinical and virological data were obtained from 129 hospitalized COVID-19 patients (89 intensive care, 40 medium care). Generalized estimating equations were used to identify if viral RNA load, detection of viral subgenomic RNA, serum neutralizing antibody response, duration of symptoms, or immunocompromised status were predictive for a positive virus culture.

**Findings:** Infectious virus shedding was detected in 23 of the 129 patients (17,8%). The median duration of shedding was 8 days post onset of symptoms (IQR 5 – 11) and the probability of detecting infectious virus dropped below 5% after 15,2 days post onset of symptoms (95% confidence interval (CI) 13,4 – 17,2). Multivariate analyses identified viral loads above 7 log_10_ RNA copies/mL (odds ratio [OR]; CI 14,7 (3,57-58,1; p<0,001) as independently associated with isolation of infectious SARS-CoV-2 from the respiratory tract. A serum neutralizing antibody titre of at least 1:20 (OR of 0,01 (CI 0,003-0,08; p<0,001) was independently associated with non-infectious SARS-CoV-2.

**Interpretation:** Infection prevention and control guidelines should take into account that patients with severe or critical COVID-19 may shed infectious virus for longer periods of time compared to what has been reported for in patients with mild COVID-19. Infectious virus shedding drops to undetectable levels below a viral RNA load threshold and once serum neutralizing antibodies are present, which warrants the use of quantitative viral RNA load assays and serological assays in test-based strategies to discontinue or de-escalate infection prevention and control precautions.

**Research in context:** *Evidence before this study:* We searched PubMed, bioRxiv, and medRxiv for articles that reported on shedding of infectious virus in COVID-19 patients using the search terms (“coronavirus” OR “SARS” OR “SARS-CoV-2” OR “COVID-19”) AND (“shedding” OR “infectivity” OR “infectious” OR “virus culture”) with no language or time restrictions. A detailed study on nine patients with mild COVID-19 reported that infectious virus could not be isolated after more than eight days of symptoms. The probability of isolating infectious virus was less than 5% when viral loads dropped below 6,51 Log_10_ RNA copies/mL. Similar results were obtained with a larger diagnostic sample set, but that study did not report on clinical parameters such as disease severity. Finally there is a report of a single patient shedding infectious virus up to 18 days after onset of symptoms. No published works were found on the shedding of infectious virus in patients with severe or critical COVID-19, and no published works were found on factors independently associated with shedding of infectious virus.

*Added value of this study:* We assessed the duration and determinants of infectious virus shedding in 129 patients with severe or critical COVID-19. The duration of infectious virus shedding ranged from 0 to 20 days post onset of symptoms (median 8 days, IQR 5 – 11). The probability of detecting infectious virus dropped below 5% after 15,2 days post onset of symptoms (95% confidence interval (CI) 13,4 – 17,2). Viral loads above 7 log_10_ RNA copies/mL were independently associated with detection of infectious SARS-CoV-2 from the respiratory tract (odds ratio [OR]; CI 14,7 (3,57-58,1; p<0,001). A serum neutralizing antibody titre of at least 1:20 (OR of 0,01 (CI 0,003-0,08; p<0,001) was independently associated with non-infectious SARS-CoV-2.

*Implications of all the available evidence:* Infection prevention and control guidelines should take into account that patients with severe or critical COVID-19 may shed infectious virus for longer periods of time compared to what has been reported for in patients with mild COVID-19. Quantitative viral RNA load assays and serological assays should be used for test-based strategies to discontinue or de-escalate infection prevention and control precautions.

## Background

Coronavirus disease 2019 (COVID-19) is a new clinical entity caused by severe acute respiratory syndrome coronavirus 2 (SARS-CoV-2). ^1,2^ In particular, persons with underlying diseases such as diabetes mellitus, hypertension, cardiovascular disease and respiratory disease are at increased risk for severe COVID-19, and case fatality rates increase steeply with age.^3^

Understanding the kinetics of infectious virus shedding in relation to potential for transmission is crucial to guide infection prevention and control strategies.^4^ Long-term shedding of viral RNA has been reported in COVID-19 patients, even after full recovery, putting serious constraints on timely discharge from the hospital or de-escalation of infection prevention and control practices.^5–7^ Detection of viral RNA by reverse transcriptase-polymerase chain reaction (RT-PCR) is the gold standard for COVID-19 diagnosis and this technique is used in test-based strategies to discontinue or de-escalate infection prevention and control precautions.^8–10^ However, there is no clear correlation between detection of viral RNA and detection of infectious virus using cell culture.^5,11,12^ Detection of infectious virus, also called “live virus” or “replication-competent virus”, by demonstration of *in vitro* infectiousness on cell lines is regarded as a more informative surrogate of viral transmission than detection of viral RNA. ^8–10^ In a COVID-19 hamster model, the window of transmission correlated well with the detection of infectious virus using cell culture but not with viral RNA.^13^ Key questions in COVID-19, like in any other infectious disease, are how long a person sheds infectious virus and what the determinants are of infectious virus shedding.^5,11,12,14,15^

Two studies reported that infectious virus could not be detected in respiratory tract samples obtained more than 8 days after onset of symptoms despite continued detection of high levels of viral RNA.^5,12^ For one patient, infectious virus shedding up to 18 days after onset of symptoms was reported.^11^

Shedding of infectious SARS-CoV-2 has not been studied in larger groups of patients nor in patients with severe or critical COVID-19. Therefore, we assessed the shedding of infectious virus in 129 severely ill COVID-19 patients using virus cultures on respiratory tract samples as a proxy, and addressed if viral RNA load in respiratory tract samples, detection of subgenomic viral RNA in respiratory tract samples, neutralizing antibody titer in serum, the duration of symptoms, and/or immunocompromised status accurately predicted infectious virus shedding.

## Methods

### Samples and Patients

Between March 8^th^ 2020 and April 8^th^ 2020, diagnostic respiratory samples of COVID-19 patients from the Erasmus MC that were send to our laboratory for SARS-CoV-2 PCR were also submitted for virus culture. From these patients, results from SARS-CoV-2 PCRs on diagnostic respiratory samples and results from SARS-CoV-2 neutralizing antibody measurements on serum samples were extracted from our diagnostic laboratory information management system. The following information was extracted from the electronic patient files: date of onset of symptoms, disease severity (hospitalized on ICU with mechanical ventilation, hospitalized on ICU with oxygen therapy, hospitalized to ward with oxygen therapy, hospitalized to ward without oxygen therapy), information to classify patients as immunocompetent, non-severely immunocompromised, or severely immunocompromised as described previously^16^, and whether the patients were still alive or not as of April 17^th^ 2020.

### Sample processing and analysis

Swabs were collected from the upper respiratory tract and sputum from the lower respiratory tract. Detailed descriptions of sample processing methods and analyses are presented in the supplementary material. In short, real-time RT-PCR detection of SARS-CoV-2 was performed using an in-house assay^17^ or using the SARS-CoV-2 test on a cobas® 6800 system (Roche Diagnostics). Subsequently, cycle threshold (ct) values were converted to Log_10_ RNA copies/mL using calibration curves based on quantified E-gene *in vitro* RNA transcripts as described before.^5^ SARS-CoV-2 subgenomic RNAs were detected with RT-PCR as described previously.^5^ Vero cells, clone 118, were used for isolation of infectious SARS-CoV-2 from respiratory tract samples.^18^ Samples were cultured for seven days, and, once cytopathic effect (CPE) was visible, the presence of SARS-CoV-2 was confirmed with immunofluorescent detection of nucleocapsid proteins. SARS-CoV-2 neutralizing antibodies were measured in serum samples using a plaque-reduction neutralization test as described previously.^19^ A plaque reduction neutralization titer 50% (PRNT50%) of 1:20 or more was considered to be positive and a PRNT50% below 1:20 negative.

### Medical ethical approval

All patient samples and data used in this study were collected in the context of routine clinical patient care. Additional analyses were performed only on surplus of patient material collected in the context of routine clinical patient care. Our institutional review board approved the use of these data and samples (METC-2015-306) and written informed consent was waived.

### Statistical analysis

Categorical and continuous variables were compared using the chi-square test the student’s t-test, respectively. Generalized estimating equations were used to identify factors that are associated with a virus culture positive respiratory tract sample. The continuous data in the generalized estimating equations were dichotomized using various cut-off values. In the main paper we present the results of the best fitting generalized estimating equations using the levels of dichotomizing that had the best fit according to the quasi-likelihood under the independence criterion (QIC).^20^ Sensitivity analysis is provided in the supplementary material. All variables having a p-value <0.1 in univariate analysis were submitted into a multivariate general estimating equation to account for repeated measurements obtained from the same patient during hospitalization.^21^ For this analysis we used the geepack package and R version 4.0.0.^21^ Probit analyses were performed with MedCalc version 19.2.3 (MedCalc Software Ltd).

## Results

We included 129 hospitalized individuals that had been diagnosed with COVID-19 by RT-PCR and for whom at least one virus culture from a respiratory tract sample was available (Table 1). Of these, 89 patients (69,0%) had been admitted to the intensive care and the remaining 40 patients (31,0%) were admitted to the medium care. Mechanical ventilation was only performed at the intensive care (81 or 91,0% of patients). Supplemental oxygen was given to 8 (9.0%) of the intensive care patients and to 35 (87,5%) medium care patients. Thirty patients were immunosuppressed (23%) of whom 19 (14,7%) were non-severely immunocompromised and 11 (8,5%) were severely immunocompromised.

**Table 1.** Patient characteristics.

| Characteristic | All | Intensive care | Medium care | p-value<br>(ICU vs ward) |
| --- | --- | --- | --- | --- |
| Number | 129 | 89 (69.0%) | 40 (31.0%) |  |
| Male | 86 (66.7%) | 65 (73.0%) | 21 (52.5%) | 0.04 |
| Age (median – IQR) | 65 (57-72) | 66 (57 – 72) | 63 (57-74) | 0.90 |
| Immunocompromised |  |  |  |  |
| Moderate | 19 (14.7%) | 10 (11.2%) | 9 (22.5%) | 0.04 |
| Severe | 11 (8.5%) | 5 (5.6%) | 6 (15.0%) |  |
| Clinical parameters |  |  |  |  |
| Mechanical ventilation | 81 (62.8%) | 81 (91.0%) | 0 |  |
| Supplemental oxygen | 43 (33.3%) | 8 (9.0%) | 35 (87.5%) |  |
| Died | 14 (10.9%) | 11 (12.3%) | 3 (7.5%) |  |
| Duration of illness* |  |  |  |  |
| Median (IQR) | 18 (13-21) | 18 (13-22) | 15 (12-18) | 0.009 |
| Tests per patient,<br>Total (mean per person) |  |  |  |  |
| Culture | 690 (5.3) | 601 (6.8) | 89 (2.2) |  |
| PRNT | 112 (0.9) | 82 (0.9) | 30 (0.8) |  |
| PCR | 688 (5.3) | 599 (6.7) | 89 (2.2) |  |

\* As of April 17<sup>th</sup> 2020. PRNT = plaque reduction neutralization titer.
Respiratory tract samples for virus culture and PCR were obtained from the lower respiratory tract (sputum) on the intensive care unit (538/690 samples, 78%) and from the upper respiratory tract (swabs) on the intensive care unit as well as on the medium care unit (152/690 samples, 22%). A total of 127 out of the 690 respiratory tract samples that were submitted for virus culture (18,4%) were obtained from immunocompromised patients.

We tested 690 respiratory samples from the 129 patients for the presence of infectious virus using cell culture and determined the viral RNA load with RT-qPCR (Figure 1). Infectious SARS-CoV-2 was isolated from 62 respiratory tract samples (9,0%) of 23 patients (17,8%). The median time of infectious virus shedding was 8 days post onset of symptoms (IQR 5 – 11, range 0 – 20) and probit analysis showed a probability of ≤ 5% for isolating infectious SARS-CoV-2 when the duration of symptoms was 15,2 days (95% CI 13,4 – 17,2) or more (Figure 2). The median viral load was significantly higher in culture positive samples than in culture negative samples (8,14 versus 5,88 Log_10_ RNA copies/mL, p<0,0001) and the probability of isolating infectious SARS-CoV-2 was less than 5% when the viral load was below 6,63 Log_10_ RNA copies/mL (95% CI 6,24 – 6,91) (Figure 2).

**Figure 1.**
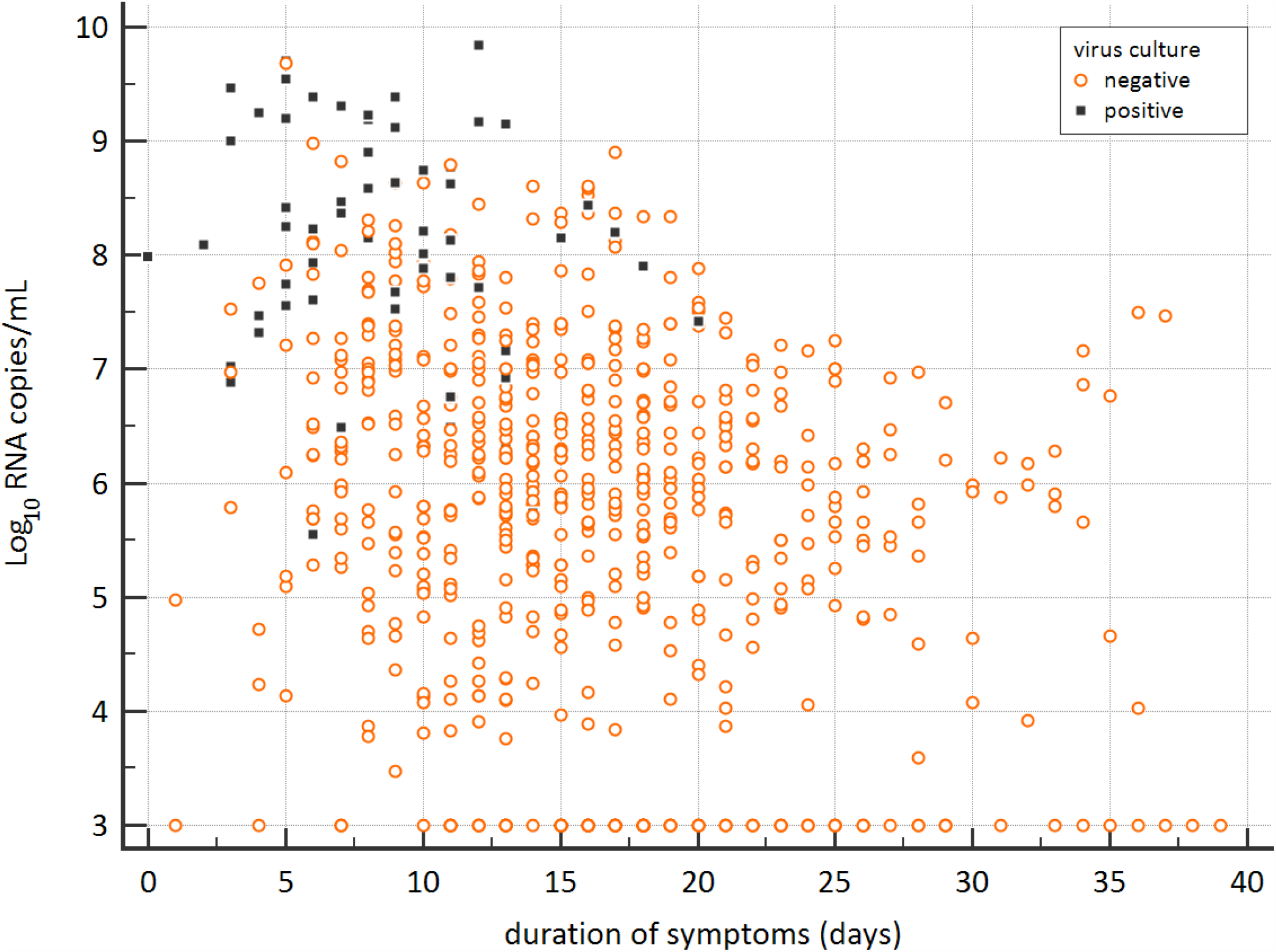
Viral RNA loads (Log_10_ RNA copies/mL) in the respiratory samples versus the duration of symptoms (days). Black boxes represent virus culture positive samples and open red circles represent the virus culture negative samples.

**Figure 2.**
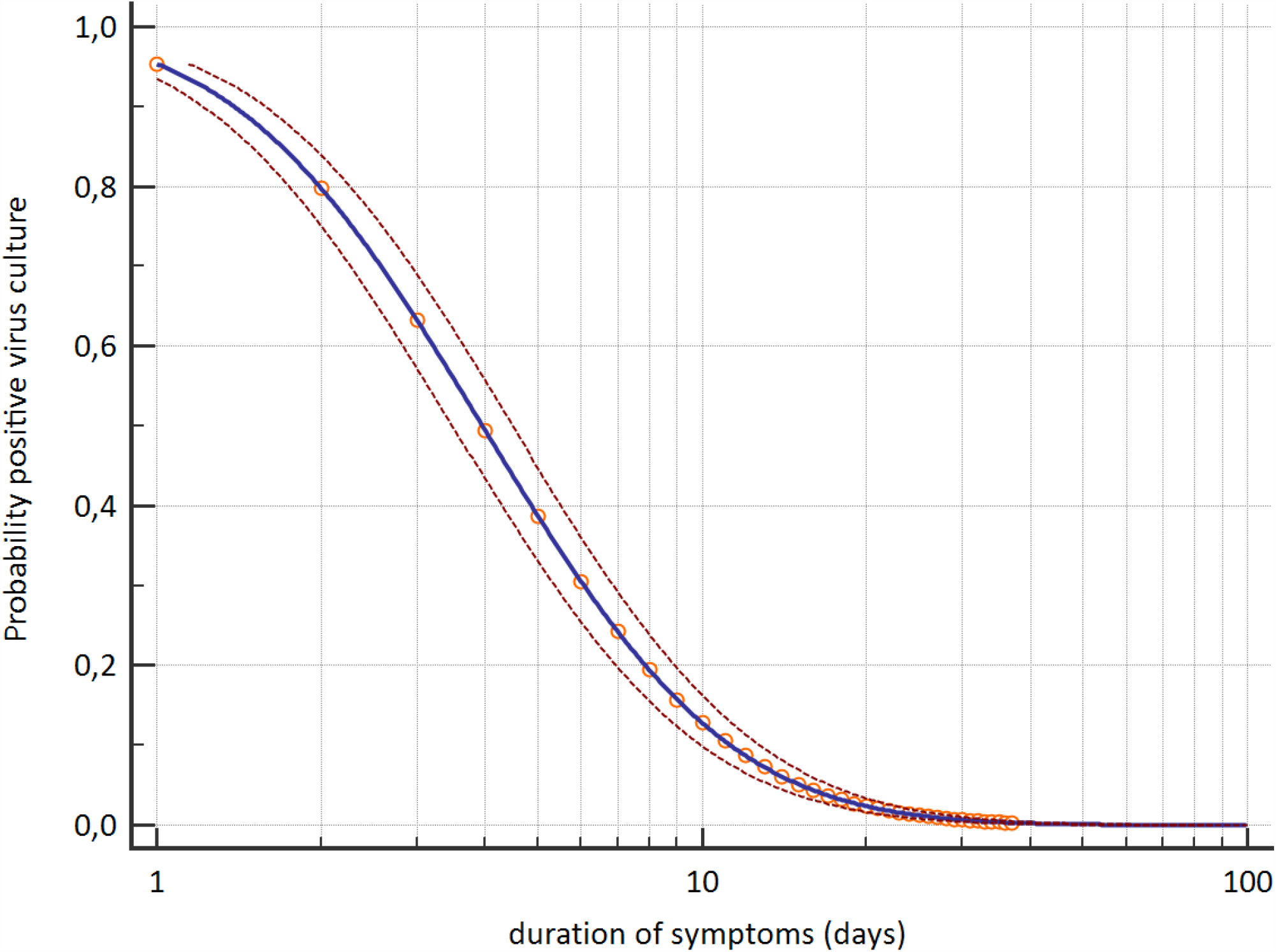
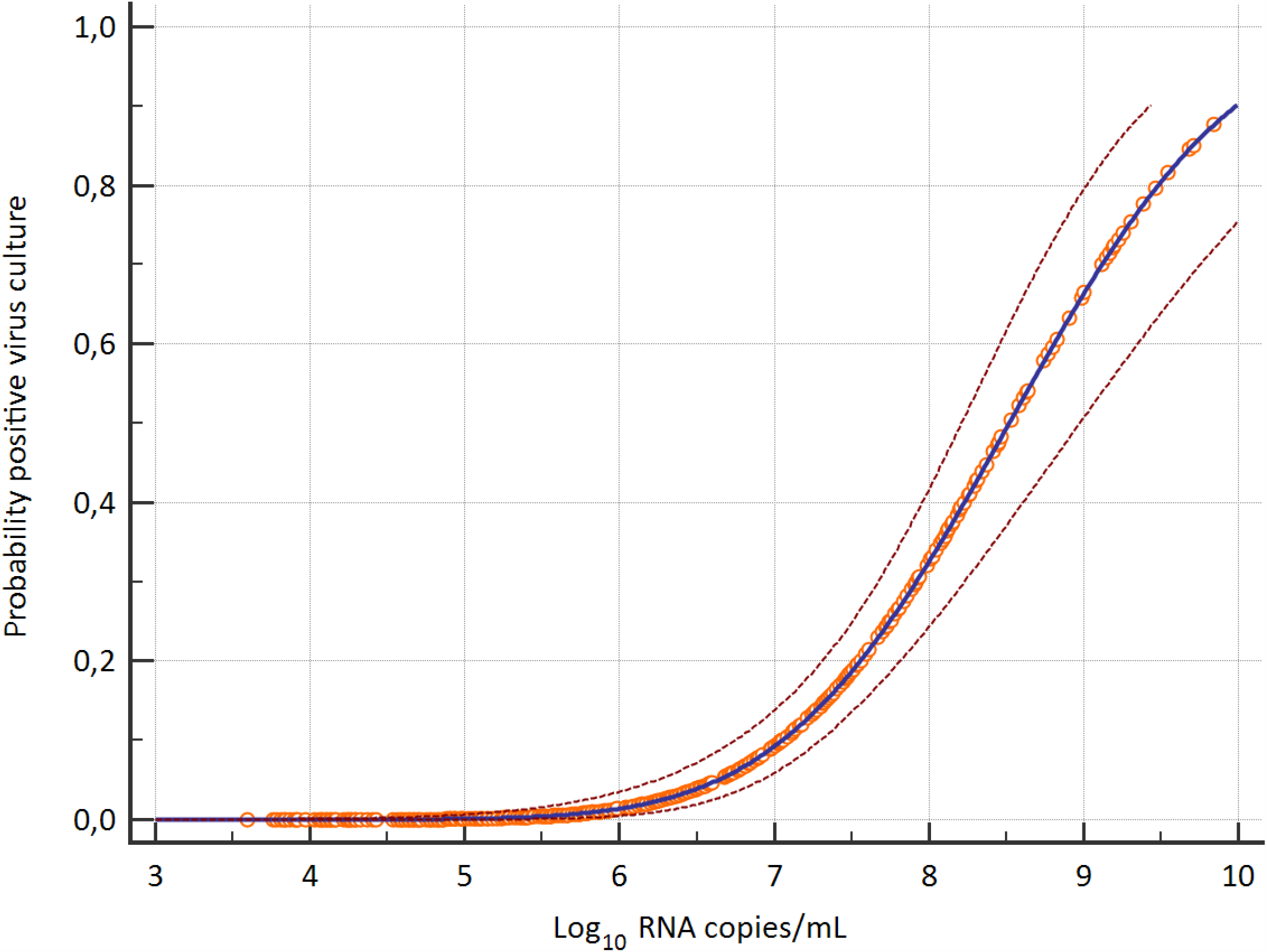
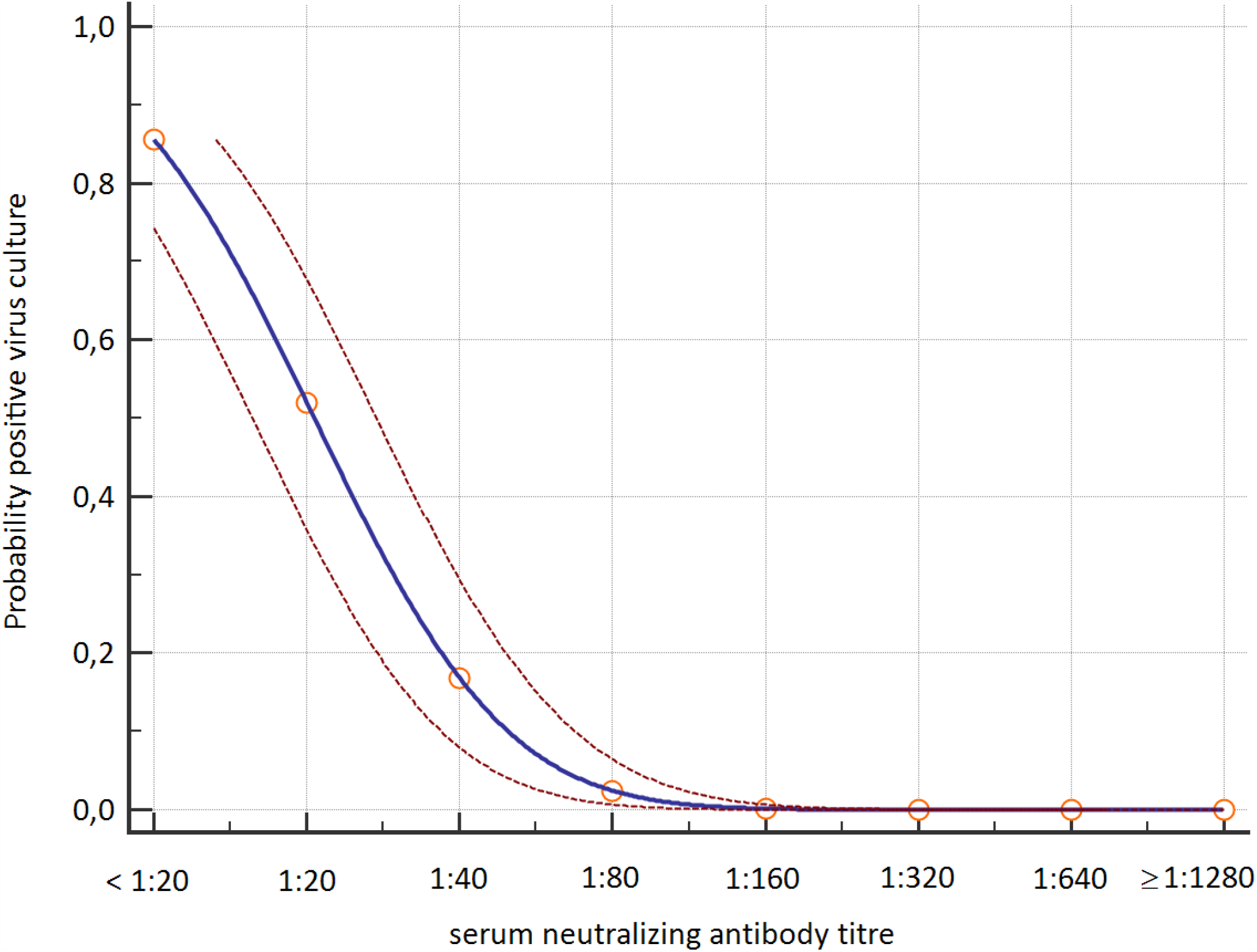
Probit analyses of the detection of infectious virus in respiratory samples with cell culture for duration of symptoms in days (upper panel), viral RNA load in Log10 copies per mL (middle panel), and serum neutralizing antibody titer (lower panel). Blue line represent the probit curve and the dotted red lines represent the 95% confidence interval. Circles are marker points. Serum neutralizing antibody titers are expressed as plaque-reduction neutralization titers 50% as described previously.^19^

For 27 patients, neutralizing antibody titers from 112 serum samples that were obtained on the same day as a respiratory tract sample were available in our diagnostic database (Table 2). The probability of isolating infectious virus was less than 5% when the neutralizing antibody titer was 1:80 or higher (Figure 2). In addition to these neutralizing antibody measurements, we performed RT-PCRs to detect SARS-CoV-2 subgenomic messenger RNA in the 112 corresponding respiratory tract samples. Detection of the subgenomic RNAs outlasted the detection of infectious virus (supplementary Figure 1), and predicted poorly if virus cultures were positive (positive predictive value of 37,5%). In addition, quantitative assessment of subgenomic RNA using cycle threshold (CT) values had no added value over measuring viral genomic RNA loads or serological response to predict infectious virus shedding (supplementary Figure 2).

**Table 2.** Serum neutralizing antibody titers and isolation of infectious virus from the respiratory tract.

| Serum neutralizing antibody titer | Total number samples | Number culture positive samples (%) | Number culture negative samples (%) |
| --- | --- | --- | --- |
| < 1:20 | 31 | 27 (87%) | 4 (13%) |
| 1:20 | 10 | 4 (40%) | 6 (60%) |
| 1:40 | 7 | 2 (29%) | 5 (71%) |
| 1:80 | 2 | 0 (0%) | 2 (100%) |
| 1:160 | 4 | 0 (0%) | 4 (100%) |
| 1:320 | 11 | 0 (0%) | 11 (100%) |
| 1:640 | 9 | 0 (0%) | 9 (100%) |
| 1:1280 | 14 | 0 (0%) | 14 (100%) |
| 1:2560 | 16 | 0 (0%) | 16 (100%) |

Serum neutralizing antibody titers against SARS-CoV-2 were determined using a plaque-reduction neutralization assay [17].
Neutralizing antibodies (titers of 1:20 or higher) were detected in 72,3% (81/112) of the serum samples. For six patients, infectious SARS-CoV-2 was isolated from the respiratory tract despite the presence of neutralizing SARS-CoV-2 antibodies in the serum sample pairs. In four of these six patients, infectious virus was not isolated in the consecutive respiratory tract samples obtained after a virus culture positive sample (sampled from day +1, +1, +4, and +4 in respect to virus culture positive sample). For one patient, infectious virus was not isolated in the respiratory tract sample obtained one day after the virus culture positive respiratory tract sample, while the respiratory tract sample obtained two days after the virus culture positive respiratory tract sample was positive for infectious SARS-CoV-2. All respiratory tract samples obtained thereafter tested negative for infectious virus. For one patient, no follow-up respiratory tract samples were available.

Finally, the key parameters were compared using multivariate generalized estimating equations (Table 3). For this, time points for which all three data types (RT-qPCR, virus culture and serum neutralizing antibody titer) were available were included (n = 112). A viral load exceeding 7 Log_10_ RNA copies/mL, less than 7 days of symptoms, absence of serum neutralizing antibodies and being immunocompromised were all associated with a positive virus culture in univariate analysis. After submitting all these variables into a multivariate analysis, we found that only a viral load above 7 Log_10_ RNA copies/mL and absence of serum neutralizing antibodies were independently associated with isolation of infectious SARS-CoV-2 from the respiratory tract.

**Table 3.** univariate and multivariate analysis of key determinants for infectious virus shedding.

| Variable | Positive virus culture<br>(n=33) | Negative virus culture<br>(n=79) | Univariate<br>Odds ratio (95% CI) | Multivariate<br>Odds ratio (95% CI) |
| --- | --- | --- | --- | --- |
| Viral RNA load<br>>10<sup>7</sup> RNA copies/mL | 29 (87.9%) | 22 (27.8%) | 18.8 (5.5 – 64.2), p<0.001 | 14.7 (3.7-58.1), p<0.001 |
| Duration of symptoms<br>< 7 days | 20 (60.6%) | 17 (21.5%) | 5.6 (1.7 – 18.1), p=0.004 | 2.1 (0.4-11.7), p=0.31 |
| Serum neutralizing antibody titer<br>1:20 or higher | 6 (18.2%) | 75 (94.9%) | 0.01 (0.003 – 0.05), p<0.001 | 0.01 (0.002-0.08), p<0.001 |
| Immunocompromised<br>yes | 10 (30.3%) | 10 (12.7%) | 3.00 (0.8-11.0), p=0.098 | 2.0 (0.7 – 5.3), p=0.22 |

Results of the univariate and multivariate generalized estimating equation analysis. The analyses were limited to the samples for which a viral RNA load and a serum neutralizing antibody titer were available from samples taken at the same day.

## Discussion

In this study we assessed the duration and key determinants of infectious SARS-CoV-2 shedding in patients with severe and critical COVID-19. Such information is critical to design test-based and symptom-based strategies to discontinue infection prevention and control precautions. Both strategies only allow for discontinuation of infection prevention and control precautions after partial resolution of symptoms. Symptom-based strategies use as additional criterion that a certain time interval should have passed since onset of symptoms, while test-based strategies use negative SARS-CoV-2 RT-PCR results as main additional criterion.

The duration of infectious virus shedding found in this study was longer than has been reported previously.^5,11,12^ Wölfel and colleagues showed for patients with mild COVID-19 that infectious virus could not be detected after more than eight days since onset of symptoms.^5^ Bullard and colleagues obtained similar results, but disease severity was not reported.^12^ Shedding of infectious virus up to 18 days after onset of symptoms has been reported for a single case of mild COVID-19.^11^ The patients in this study had severe or critical COVID-19 and detection of infectious virus was common after eight days or more since onset of symptoms. For a single patient, infectious virus was detected up to 20 days after onset of symptoms. Higher viral loads have been reported for severe COVID-19 cases compared to mild cases, which may in part explain the longer duration of shedding found in this study.^22–26^ Our findings imply that symptom-based strategies to discontinue infection prevention and control precautions should take diseases severity into account. For example, the CDC currently use a minimum disease duration of 10 days in their symptom-based strategy as the statistically estimated likelihood of recovering replication-competent virus approaches zero after ten days of symptoms.^8,27^ Based on our findings, a longer disease duration could be considered for severely ill patients.

High viral RNA loads were independently associated with shedding of infectious virus, but, upon seroconversion, shedding of infectious virus dropped rapidly to undetectable levels. Infectious virus could not be isolated from respiratory tract samples once patients had a serum neutralizing antibody titer of at least 1:80. These results warrant the use of quantitative viral RNA load assays and serological assays in test-based strategies to discontinue or de-escalate infection prevention and control precautions. The probability of isolating infectious virus was less than 5% when viral RNA load was below 6,63 Log10 RNA copies/mL, which is strikingly similar compared to the cut-off of 6,51 Log10 RNA copies/mL reported by Wölfel and colleagues.^5^ In addition, Bullard and colleagues used cycle threshold (ct) values as quantitative measure for viral RNA load and reported that infectious virus could not be isolated from diagnostic samples when ct values were above 24.^12^ Together, these results indicate that viral RNA load cut-offs could be used in test-based strategies to discontinue infection prevention and control precautions. In addition, we report here a very strong association between neutralizing antibody response and shedding of infectious virus with an odds ratio of 0,01 for isolating infectious virus after seroconversion. Antibody responses were measured with a plaque-reduction neutralization test (PRNT).^19^ Neutralization assays, which are the gold standard in coronavirus serology, are labor-intensive and require a biosafety level 3 laboratory. Some commercial immunoassays, which require less stringent biosafety measures and are amenable to high throughput use, have shown good agreement with PRNT.^28^ In particular immunoassays that correlate well with serum neutralization titers of 1:80 or higher would be useful for test-based strategies as we did not detect infectious virus shedding in patients with these titers.

Detection of viral subgenomic RNA correlated poorly with shedding of infectious virus. These RNAs are produced only in actively infected cells and are not packaged into virions. Subgenomic RNAs were still detected when virus cultures turned negative. This could indicate that active replication continues in severely-ill symptomatic COVID-19 patients after seroconversion and after shedding of infectious virus has stopped. Possibly, infectious virions are produced but are directly neutralized by antibodies in the respiratory tract. On the other hand, the half-life of viral subgenomic RNAs is not known in COVID-19 and these RNAs may still be detected once replication has stopped.

Our study has some limitations. Firstly, virological data was obtained from diagnostic samples only and samples were not prospectively collected at predefined timepoints. However, as many aspects of COVID-19 were still unclear, a sampling-rich diagnostic approach was applied in our institution with regular virological monitoring of confirmed COVID-19 patients. This approach resulted in a large high quality dataset from a considerable number of patients including patients with a immunocompromised status. The strikingly similar viral RNA load cut-off for a 5% probability of a positive virus culture found by us and by Wölfel and colleagues underpins the validity of the results.^5^ Secondly, we used *in vitro* cell cultures as a surrogate marker for infectious virus shedding. The success of SARS-CoV-2 isolation is dependent on which cell lines is used.^29^ Vero cells are currently regarded as the gold standard to detect infectious SAR-CoV-2, but the true limit of detection is unknown. Notwithstanding the above, experimental evidence from a COVID-19 hamster model showed that transmission of SARS-CoV-2 correlated well with detection of infectious SARS-CoV-2 from respiratory tract samples using *in vitro* Vero cell cultures while detection of viral RNA did not.^13^ More data from experimental models, and epidemiological and modeling studies on transmission which take viral RNA load and antibody response into account are needed for further validation of this approach. It should be noted that, besides the infectious viral load, additional factors determine virus transmissibility. Finally, our study only included hospitalized symptomatic adults with severe or critical COVID-19 and important differences were noted in our study compared to what has been reported for in mild COVID-19. Thus, further studies are needed on the determinants and duration of infectious virus shedding in specific patient groups.

In conclusion, infection prevention and control guidelines should take into account that patients with severe or critical COVID-19 may shed infectious virus for longer periods of time compared to what has been reported for in patients with mild COVID-19. Infectious virus shedding drops to undetectable levels when viral RNA load is low and serum neutralizing antibodies are present, which warrants the use of quantitative viral RNA load assays and serological assays in test-based strategies to discontinue or de-escalate infection prevention and control precautions.

## Data Availability

Data available upon request.

## Acknowledgements

We gratefully acknowledge EVA-g and Christian Drosten for provision of the quantified E-gene transcript. Ga-Lai Chong, Rose Willemze, Jordy Dekker, George Sips, Stephanie Popping, Daphne Mulders, Alex de Ries and Jeroen Ijpelaar gratefully acknowledged for their technical and analytical contributions. This work partially was funded through EU COVID-19 grant RECOVER 101003589.

## Figure legends

**Supplementary Figure 1A.**
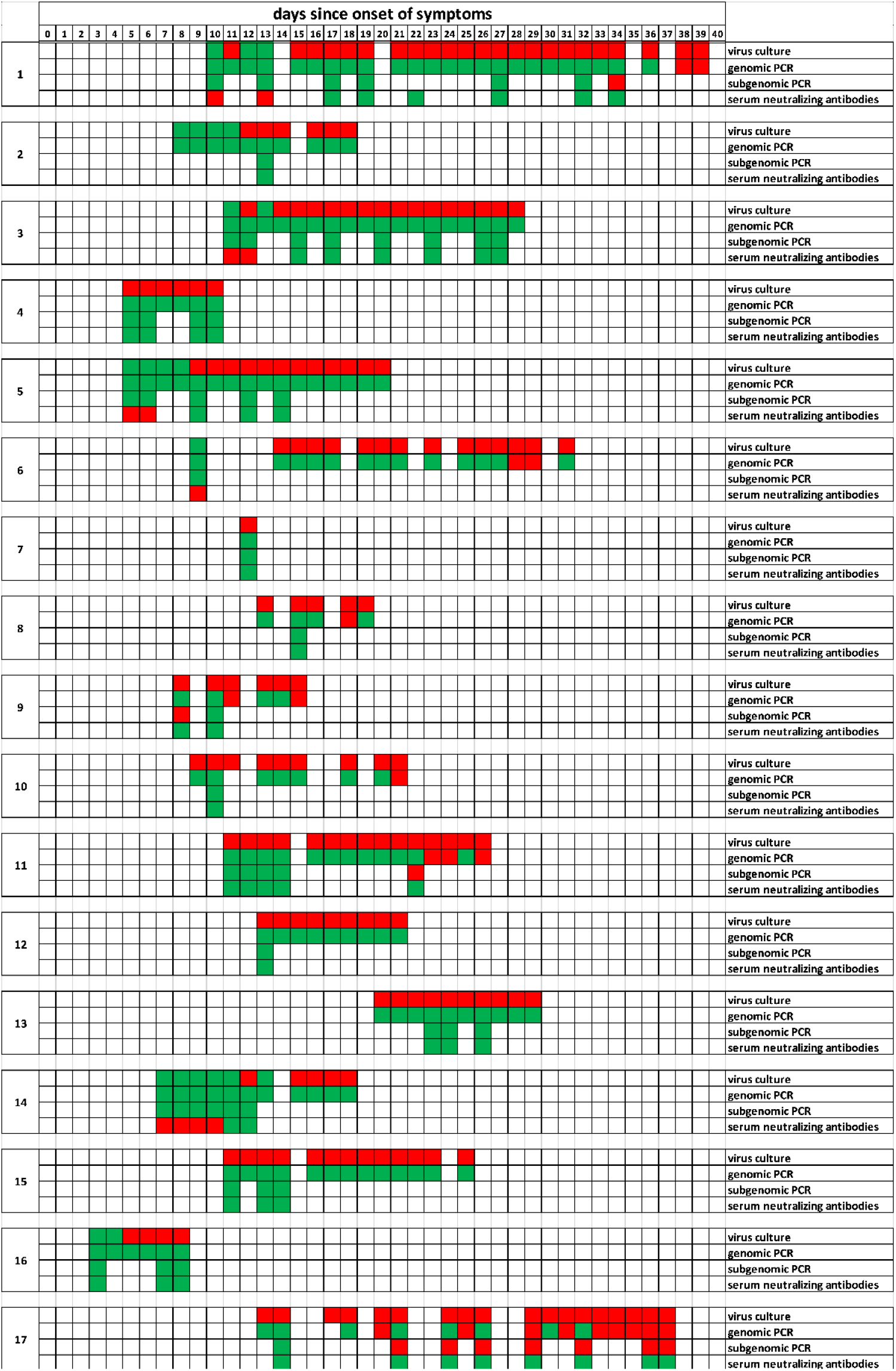
Qualitative assessment of sgRNA in lower respiratory tract samples. Individual patient charts with virological test results (positive/negative) in relation to the duration of symptoms for patients for whom sgRNA RT-PCR results of lower respiratory tract samples were available. Positive test results are depicted in green and negative test results in red.

**Supplementary Figure 1B.**
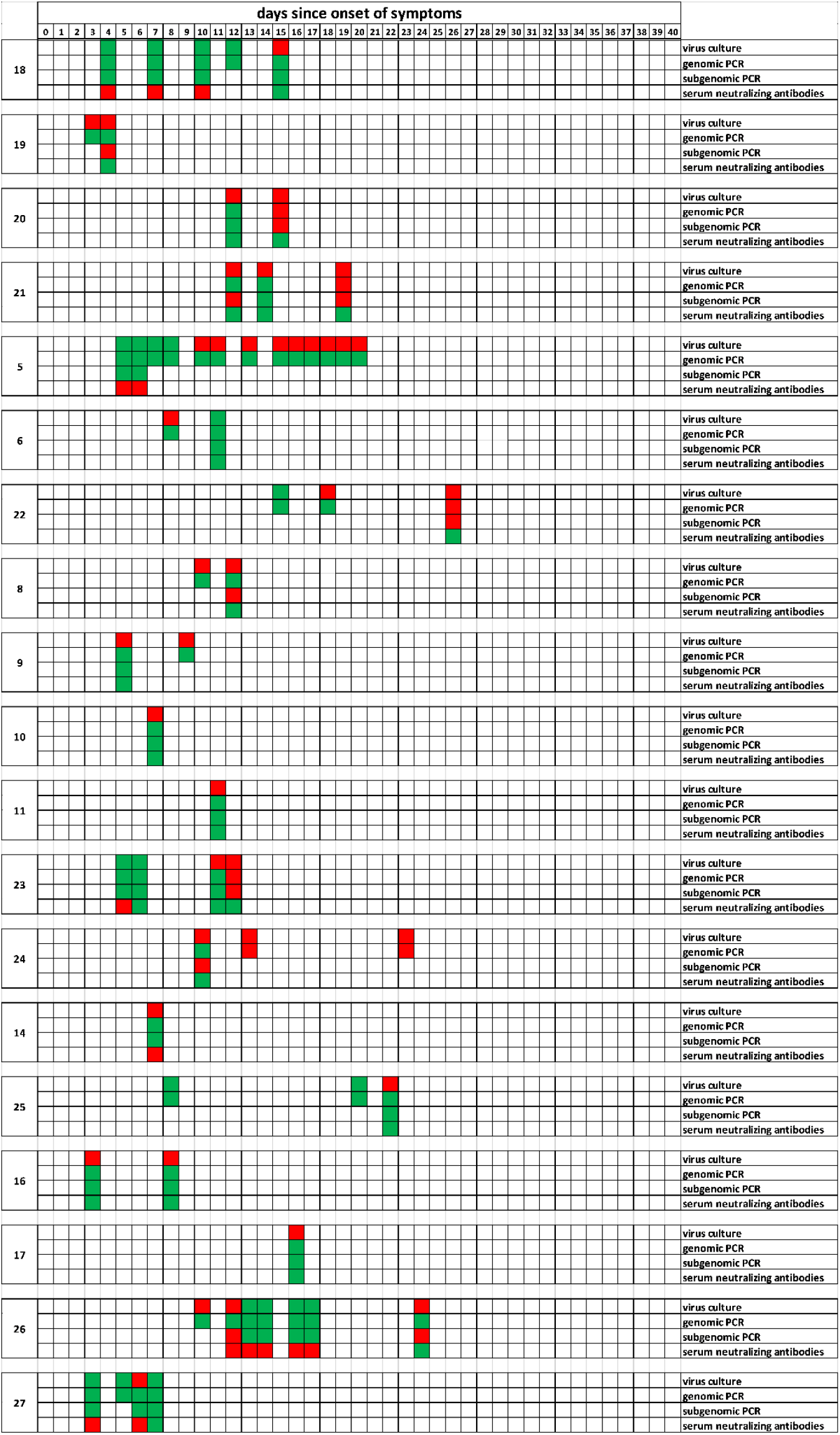
Qualitative assessment of sgRNA in upper respiratory tract samples. Individual patient charts with virological test results (positive/negative) in relation to the duration of symptoms for patients for whom sgRNA RT-PCR results of upper respiratory tract samples were available. Positive test results are depicted in green and negative test results in red.

**Supplementary Figure 2.**
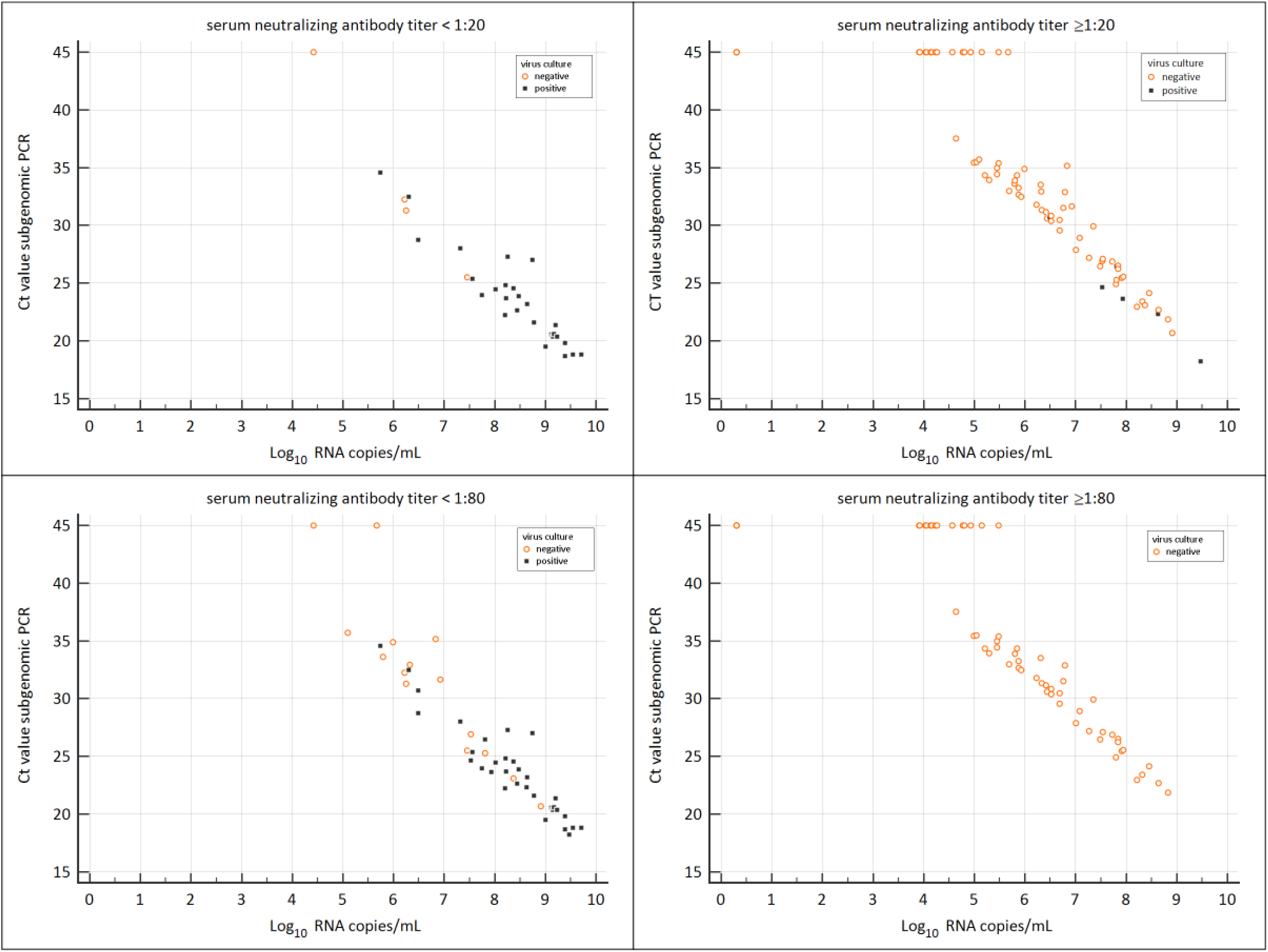
Quantitative assessment of sgRNA in respiratory tract samples. Plots of cycle threshold value of the viral subgenomic RNA RT-PCR (y-axis) versus the genomic viral load (x-axis) for patients with an undetectable neutralizing antibody response (upper left panel), a detectable neutralizing antibody response (upper right panel), neutralizing antibody titers < 1:80 (lower left panel), and neutralizing antibody titers ≥ 1:80. Cycle threshold values are inversely correlated to the subgenomic RNA load and a cycle threshold > 45 is regarded as a negative test result. Ct values of the subgenomic RNA correlated well with genomic viral RNA load, but had no added value over this measure or antibody response to predict a positive virus culture.

## Supplement

### Processing of respiratory samples

Swabs from the upper respiratory tract were collected in tubes containing 4 mL virus transport medium (Dulbecco’s modified eagle’s medium (DMEM, Lonza) supplemented with 40% FBS, 20 mM 4-(2-hydroxyethyl)-1-piperazineethanesulfonic acid (HEPES), NaCO3, 10 microgram/ml amphotericin B, 1000 U/mL penicillin, 1000 microgram/mL streptomycin). Supernatant was passed through a 45 micrometer filter and used for PCR analysis and virus culture. For sputum samples, 6 mL sample processing medium (DMEM supplemented with 17 mM HEPES, NaCO3, 1000 U/mL penicillin, 1000 microgram/mL streptomycin, 12.5 microgram/ml amphotericin B) was added until the final volume was 6 mL. Subsequently, samples were vortexed, centrifugated, passed through a 45 micrometer filter, and 1 part FBS was added to 1.5 parts supernatant. Subsequently, processed samples were used for PCR analysis and virus culture.

### SARS-CoV-2 virus culture

Respiratory samples were cultured on Vero cells, clone 118, using 24-wells plates with glass coverslips. Cells were inoculated with 200 microliter sample per well and centrifugated for 15 minutes at 3500 g. After centrifugation, inoculum was discarded, virus culture medium (Iscove’s modified Dulbecco’s medium (IMDM; Lonza) supplemented with 2mM L-glutamine (Lonza), 100 U/mL penicillin (Lonza), 100 ug/mL streptomycin (Lonza), 2.5 microgram/mL amphotericin B (department of hospital pharmacy, Erasmus MC), and 1% heat-inactivated fetal bovine serum (Sigma)) was added, and samples were cultured at 37 °C and 5% CO_2_ for 7 days. Each sample was cultured in triplicate: Two replicates were fixed with ice-cold acetone after 24 hours and 48 hours respectively irrespective if cytopathic effect (CPE) was visible. The fixed samples were further analyzed with immunofluorescence (see below). The remaining replicate was scored for CPE on a daily basis for 7 days. When CPE was visible, the sample was fixed with ice-cold acetone and further analyzed with immunofluorescence (see below). Virus cultures were regarded as negative if no CPE was visible during 7 days. For immunofluorescence read-out, the fixed cells were washed with phosphate buffer saline (PBS), and incubated for 30 minutes at 37 °C with 25 microliters 1000-fold diluted polyclonal rabbit SARS-CoV anti-nucleoprotein antibodies (Sino Biological, catalogue number 40143-T62). After incubation, samples were washed with three times with PBS and once with deionized water. Subsequently, cells were incubated for 30 minutes at 37 °C with 25 microliters 2000-fold diluted Alexa Fluor 488 - labeled polyclonal goat anti-rabbit IgG (Invitrogen, catalogue number A-11070). Subsequently, cells were washed three times with PBS. Finally, cells were incubated for 1 minute with 25 microliters Evan’s Blue (counterstain), washed twice with deionized water, air dried and analyzed with a fluorescence microscope.

### Statistical analysis

For the generalized estimating equations, we only included the 112 respiratory tract samples (33 with a positive culture, 79 with a negative culture) for which serum neutralizing antibody titer results obtained on the same day were available. The continuous data in the generalized estimating equations were dichotomized in the main analysis. In Table S1 and S2 (see below), we present the results of a sensitivity analysis in which we show that choosing a different cut-off value for dichotomizing the different dependent variables did not have an impact on which of these variables had a statistically significant and independent impact on a positive culture and therefore of finding an infectious virus. For each of the cut-offs we also calculated the quasi-likelihood under the independence model criterion (QIC) as a criterion to identify the best fitting generalized estimating equation model. The statistical model with the lowest QIC values, indicating the best fit, has been presented in the main paper.

**Table S1.** Results of the univariate sensitivity analysis.

| Characteristic | Culture positive<br>(n=33) | Culture negative<br>(n=79) | Odds ratio (95%<br>confidence interval) | p | QIC |
| --- | --- | --- | --- | --- | --- |
| Viral load (RNA copies/mL) |  |  |  |  |  |
| > 7 log<sub>10</sub> | 29 (88%) | 22 (29%) | 18.8 (5.5- 64.2) | <0.001 | 105.3 |
| > 8 log<sub>10</sub> | 23 (70%) | 7 (9%) | 23.7 (9.3- 60.4) | <0.001 | 98.3 |
| Duration of illness |  |  |  |  |  |
| < 7 days | 12 (36%) | 6 (8%) | 7.0 (1.5-32.7) | 0.01 | 131.3 |
| <10 days | 20 (61%) | 17 (22%) | 5.6 (1.7-18.1) | 0.004 | 128.0 |
| <14 days | 30 (91%) | 41 (52%) | 9.3 (1.1-80.3) | 0.04 | 128.3 |
| PRNT response |  |  |  |  |  |
| Detectable ≥1:20 | 6 (18%) | 75 (95%) | 0.01 (0.003-0.05) | <0.001 | 69.9 |
| Immunosuppression |  |  |  |  |  |
| Mild/moderate | 10 (30%) | 10 (13%) | 3.0 (0.8-11.0) | 0.098 | 139.1 |
| Severe | 4 (12%) | 6 (18%) | 1.7 (0.3-9.7) | 0.57 | 143.5 |

**Table S2.** sensitivity analysis of the multivariate analysis.

| Characteristic | Cut-off | Odds ratio (95% confidence interval) | p | QIC |
| --- | --- | --- | --- | --- |
| Viral load | > 7 log<sub>10</sub> | 14.7 (3.7-58.1) | <0.001 | 57.6 |
| Duration of illness | < 7 days | 2.1 (0.4-11.8) | 0.40 |  |
| PRNT | Detectable (1:20) | 0.01 (0.003-0.08) | <0.001 |  |
| Immunosuppression | yes | 2 (0.7-5.3) | 0.17 |  |
| Viral load | > 7 log<sub>10</sub> | 14.3 (3.1-65.6) | <0.001 | 61.5 |
| Duration of illness | < 10 days | 2.5 (0.5-11.7) | 0.26 |  |
| PRNT | Detectable (1:20) | 0.01 (0.002-0.09) | <0.001 |  |
| Immunosuppression | yes | 1.5 (0.5-4.9) | 0.49 |  |
| Viral load | > 7 log<sub>10</sub> | 14.8 (3.5-62.4) | <0.001 | 58.5 |
| Duration of illness | < 14 days | 2.3 (0.4-13.4) | 0.37 |  |
| PRNT | Detectable (1:20) | 0.01 (0.002-0.09) | <0.001 |  |
| Immunosuppression | yes | 1.6 (0.6-4.6) | 0.35 |  |
| Viral load | > 8 log<sub>10</sub> | 10.7 (3.7-30.8) | <0.001 | 61.4 |
| Duration of illness | < 7 days | 5 (0.6-42.6) | 0.14 |  |
| PRNT | Detectable (1:20) | 0.03 (0.006-0.13) | <0.001 |  |
| Immunosuppression | yes | 1.7 (0.5-5.8) | 0.40 |  |
| Viral load | > 8 log<sub>10</sub> | 8.6 (3-25) | <0.001 | 65.3 |
| Duration of illness | < 10 days | 3.1 (0.5-20.5) | 0.24 |  |
| PRNT | Detectable (1:20) | 0.02 (0.005-0.12) | <0.001 |  |
| Immunosuppression | yes | 1.2 (0.3-5.5) | 0.82 |  |
| Viral load | > 8 log<sub>10</sub> | 9.8 (3.7-26.0) | <0.001 | 63.4 |
| Duration of illness | < 14 days | 4.5 (0.7-28.0) | 0.11 |  |
| PRNT | Detectable (1:20) | 0.03 (0.006-0.12) | <0.001 |  |
| Immunosuppression | yes | 1.3 (0.4-4.0) | 0.70 |  |

